# Identification of endothelial-derived proteins in plasma associated with cardiovascular risk factors

**DOI:** 10.1101/2021.02.08.21251209

**Authors:** MJ Iglesias, LD Kruse, L Sanchez-Rivera, L Enge, P Dusart, MG Hong, M Uhlén, T Renné, JM Schwenk, G Bergstrom, J Odeberg, LM Butler

## Abstract

Endothelial cell (EC) dysfunction is a well-established response to cardiovascular disease (CVD) risk factors, such as smoking and obesity. Risk factor exposure can modify EC signalling and behaviour, leading to arterial and venous disease development. Biomarker panels to assess EC dysfunction are lacking, but could be useful for risk stratification and to monitor treatment response. Here, we used affinity proteomics to identify EC-derived proteins circulating in plasma that were associated with CVD risk factor exposure. 216 proteins, known to be expressed in ECs across vascular beds, were measured in plasma samples (n=1005) from the population-based *Swedish CArdioPulmonary bioImage Study* (SCAPIS) pilot. We identified 38 EC-derived proteins that were associated with body mass index, total cholesterol, low density lipoprotein, smoking, hypertension or diabetes. Sex-specific analysis revealed female- and male-only associations were most frequently observed with BMI, or total cholesterol, respectively. We showed a relationship between individual CVD risk, calculated with the Framingham risk score, and the corresponding biomarker profiles; presenting the concept of measuring EC-derived proteins in plasma to infer vascular status.

## INTRODUCTION

Cardiovascular disease (CVD) is the leading cause of death, killing approximately 17.9 million people each year, globally (WHO, 2019). Endothelial cells (EC), which line the inside of all blood vessels, are key for vascular health as they regulate haemostasis, provide an anti-thrombotic surface, control inflammation, vascular tone, angiogenesis and the transport of molecules and nutrients to and from the blood stream (1, 2). Disruption in this normal function is termed ‘EC dysfunction’, which is linked to thrombosis formation, uncontrolled leukocyte recruitment and platelet activation, inappropriate vasoconstriction and impaired recovery from injury (3). Exposure to CVD risk factors, such as smoking, obesity, hypertension, modified blood lipid profile or diabetes can induce such changes, through the effects of chronic inflammation, oxidative stress, local hypoxia and disruption in laminar flow (4-8). These changes can predispose to arterial (9, 10) and venous (11, 12) CVD development, whilst interventions which improve EC function can lessen these effects (3, 13-15). Indeed, some successful therapies for CVD have transpired to act, at least partly, through EC protective effects (e.g. angiotensin-converting enzyme inhibitors and statins), although they were not originally designed to function via this mechanism (15, 16).

Vasoreactivity has been used to assess EC dysfunction, but it can be invasive and time consuming to measure, and its prognostic value is debated (13). Plasma levels of the inflammatory marker c-reactive protein (CRP) has been suggested as a proxy for EC dysfunction, due to proposed deleterious effects on the production of the vasoprotective agent nitric oxide (NO) (3, 17, 18), although other studies dispute this (19). Plasma asymmetrical dimethylarginine (ADMA), symmetrical dimethylarginine (SDMA) and homoarginine (agents involved in NO synthesis) can be modified by exposure to CVD risk factors and have also been measured to infer EC status (20-23). Other inflammation-related markers, such as interleukin-6, intercellular adhesion molecule-1 (ICAM1), vascular adhesion molecule-1 (VCAM1), E-selectin, P-selectin and von Willebrand factor (VWF) have been used as markers for EC dysfunction (23-26). Plasma levels of these markers can be increased in response to CVD risk factor exposure (27-29) and higher E- and P-selectin levels were associated with impaired acetylcholine-dependent EC vasodilation in humans (30), indicating that they can be linked to vascular changes beyond the inflammatory response. It remains unclear whether EC dysfunction is synonymous with, or limited to, inhibited vasoreactivity or inflammation-induced activation. Furthermore, of the aforementioned markers, only E-selectin and VCAM1 have high EC-specificity (and then only under conditions of inflammation); other markers used can originate from elsewhere, e.g. plasma P-selectin is predominantly from platelets, rather than EC (31), making interpretation in the context of vascular health more complex. Sex is known to have an important influence on CVD development (32); risk factors, incidence, age of occurrence, severity, clinical presentation and treatment response varies between males and females (33-35). Despite this, sex-specific biomarkers for vascular dysfunction are lacking. Thus, the scope and specificity of the current measurements of EC dysfunction is limited (36) and we lack clinical tools for risk profiling and monitoring of response following intervention to improve EC function.

Here, we used affinity proteomics to measure levels of 216 plasma proteins, which we previously identified as having a high level of EC-specific expression across human vascular beds (37), in samples collected as part of the population-based study, the *Swedish CArdioPulmonary bioImage Study* (SCAPIS) pilot (38) (n=1005). We identified 38 EC-derived proteins in plasma that were associated with exposure to CVD risk factors. Nine proteins were associated with three risk factors or more, including ERG, VWF and HEG1. Sex specific analysis revealed that female- or male-only associations were predominantly observed between EC-derived protein levels and BMI or cholesterol, respectively. Finally, we show a relationship between biomarker expression and CVD risk, determined by the Framingham Score, presenting the concept of risk prediction through measurement of EC-derived proteins in the plasma.

## RESULTS

In order to identify potential biomarkers for endothelial (EC) dysfunction, we generated a protein candidate list for screening, based on our previous work, where we identified 234 protein coding transcripts that were endothelial enriched in human organs (i.e., with significantly higher specificity to EC, vs. other cell types) (37). Antibodies targeting 216 of the proteins encoded by these transcripts were selected from the Human Protein Atlas (HPA) project (www.proteinatlas.org/) based on availability, concentration and reliability score (Figure S1 and Table S1, Tab_2 [column A-B]). We performed affinity proteomic screening of plasma samples collected as part of the population-based *Swedish CArdioPulmonary bioImage Study* (SCAPIS) pilot, where subjects aged 50-64 years were randomly selected from the Swedish population register (38) (n=1005 [female n=507, male n=498]) (study population details Table S1, Tab_1. Five protein candidates were excluded from subsequent analysis as the reported data was affected by sampling location, indicating a stability issue and hence reduced suitability as possible candidates for a routine diagnostic setting (Figure S1 and Table S1, Tab_2 marked red).

### EC-derived plasma proteins are associated with CVD risk factors

We identified 38 EC-derived candidate proteins (18% of all tested) that were associated with one or more of the following risk factors in both sexes: body mass index (BMI), total cholesterol (CHO), low density lipoprotein (LDL), hypertension (HYP), smoking (SMK) or diabetes (DIAB). A total of 21 proteins were associated with a single CVD risk factor (Figure 1A, red and green coloured boxes represent positive and negative association, respectively, Table S1 Tab_2), whilst 17 were associated with two or more risk factors (Figure 1A, grey circles show number of proteins linked to all connecting risk factors and Figure 1B). When associated with multiple risk factors, proteins were either consistently elevated with increased risk, e.g., ERG (Figure 1B, red box); detected at higher levels with increasing BMI, LDL, smoking and hypertension (+ sign), or consistently reduced with elevated risk e.g. HEG1 (Figure 1B, green box); detected at lower levels with increasing BMI, LDL and hypertension (-sign).

**Figure 1.**
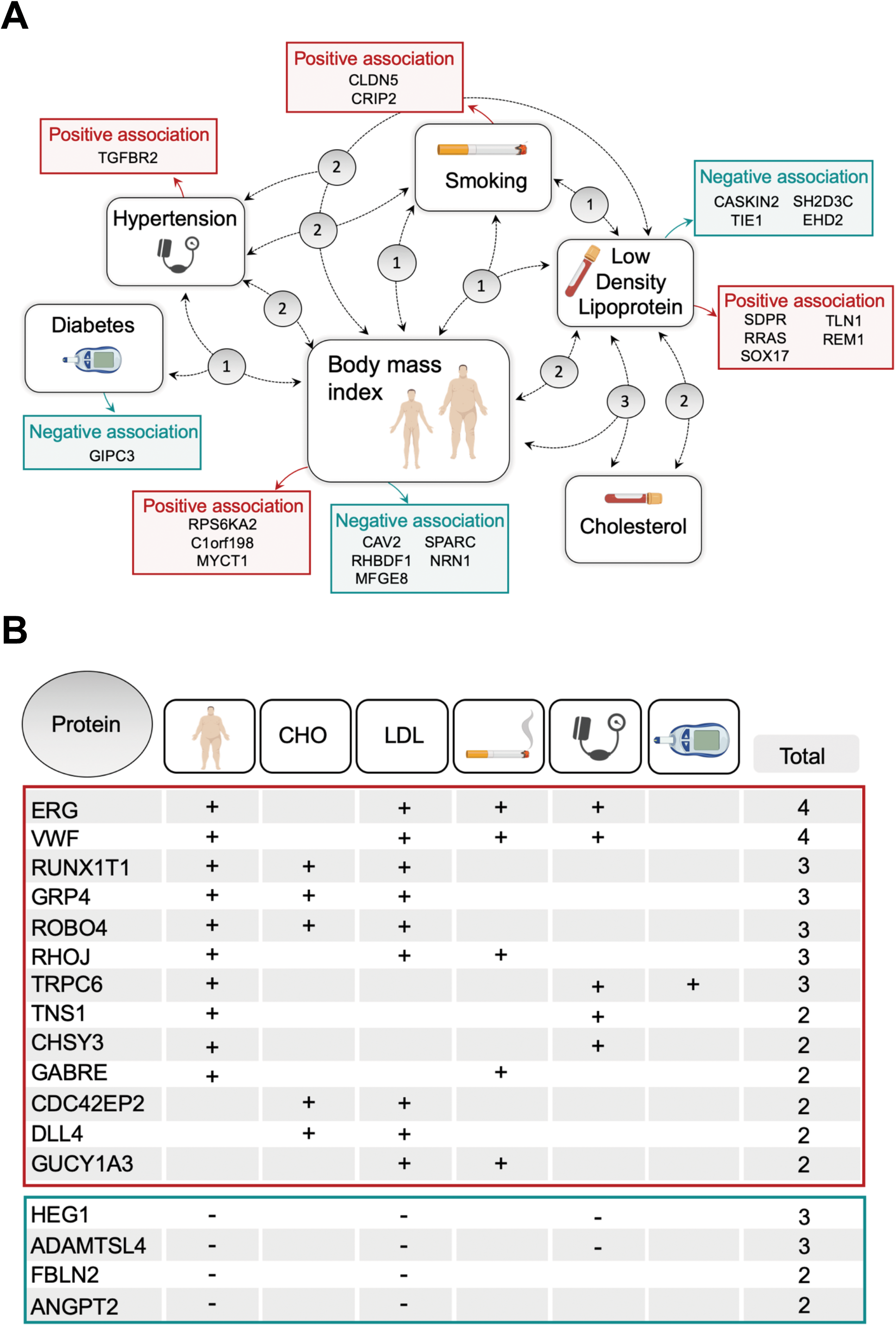
Endothelial-derived plasma protein levels are associated with cardiovascular disease (CVD) risk factors. 216 endothelial (EC)-enriched proteins were measured in plasma samples from male (n=498) and female (n=507) participants in the *Swedish CArdioPulmonary bioImage Study* (SCAPIS) pilot. Candidates associated with the CVD risk factors body mass index (BMI), total cholesterol (CHO), low density lipoprotein (LDL), smoking (SMK), hypertension (HYP) or diabetes (DIAB) were identified (age and sex adjusted linear model [Bonferroni corrected]). (**A**) Summary of EC-derived plasma proteins associated with CVD risk factors. Proteins associated with a *single* risk factor are displayed in adjacent red or green boxes, indicating a positive association (i.e. higher protein levels associated with *increased* risk, e.g. higher BMI, smoking) or a negative association (i.e. lower protein levels associated with *reduced* risk profile, e.g. lower BMI, lower blood LDL), respectively. Grey bubbles show the number of proteins associated with the (multiple) risk factors linked by the corresponding dotted line. Table (**B**) provides further details of these multiple-risk factor associated proteins, indicating if they are positively (+) or negatively associated (−) with the higher risk profile.

Obesity is associated with increased risk of both arterial and venous CVD (39, 40), with BMI being an independent predictor of disease occurrence, even after adjustment for other risk factors (40, 41). We identified 22 EC-derived proteins that were associated with BMI, making it the risk factor most frequently associated with modified levels of EC-derived plasma proteins (Figure 1A and B). 20/22 (91%) of the BMI-associated EC-derived plasma proteins were also associated with CRP, with the same effect direction (Table S1, Tab_2). CHO and diabetes were associated with plasma levels of five and two EC-derived protein(s), respectively, representing the risk factors associated with the lowest number of proteins.

### EC-derived plasma proteins can be associated with multiple risk factors

We identified a set of 17 EC-derived proteins that were positively (n=13), or negatively (n=4) associated with two or more CVD risk factors, in both sexes (Figure 1B). There were 10 EC-derived proteins associated with both BMI and LDL, making them the most common shared risk factors (Figure 1B). Consistent with previous reports (42), there was no correlation between LDL and BMI across the cohort (rho <0.1) and adjustment for BMI did not affect any EC-derived protein associations with LDL, or vice versa, further indicating that these risk factor associations were independent. 7/22 (32%) of the BMI-associated proteins were also associated with hypertension. Elevated BMI is well known to be linked to hypertension (43, 44), and the hypertension group (n=317), on average, had slightly higher BMI than the non-hypertension group (n=688) (mean ± standard dev.: 26.5±4.0 vs. 29.2±4.8 p<0.00001). For proteins associated with both, adjustment for the other factor reduced association strength, consistent with a probable interplay. EC-derived protein associations with smoking were not modified by adjustment for any other risk factor.

Representative expression plots are shown for selected proteins associated with multiple risk factors (in the full cohort [’All’], male only [’Male’] and female only [’Female’] samples) (Figure 2A-C). GRP4 was positively associated with BMI (categorised into normal, over weight or obese) (Figure 2A.i), LDL (Figure 2 A.ii) and CHO (Figure 2A.iii) (both categorised as very low, low, moderate, high or very high). TRCP6 was also positively associated with BMI (Figure 2 B.i), however, in contrast to GPR4, levels were not associated with LDL or CHO, but were associated with hypertension (Figure 2B.ii) and diabetes (Figure 2B.iii). HEG1 was negatively associated with BMI (Figure 2C.i), LDL (Figure 2C.ii) and hypertension (Figure 2C.iii). Thus, risk factor associations with EC-derived protein level could be both protein- and risk factor-specific.

**Figure 2.**
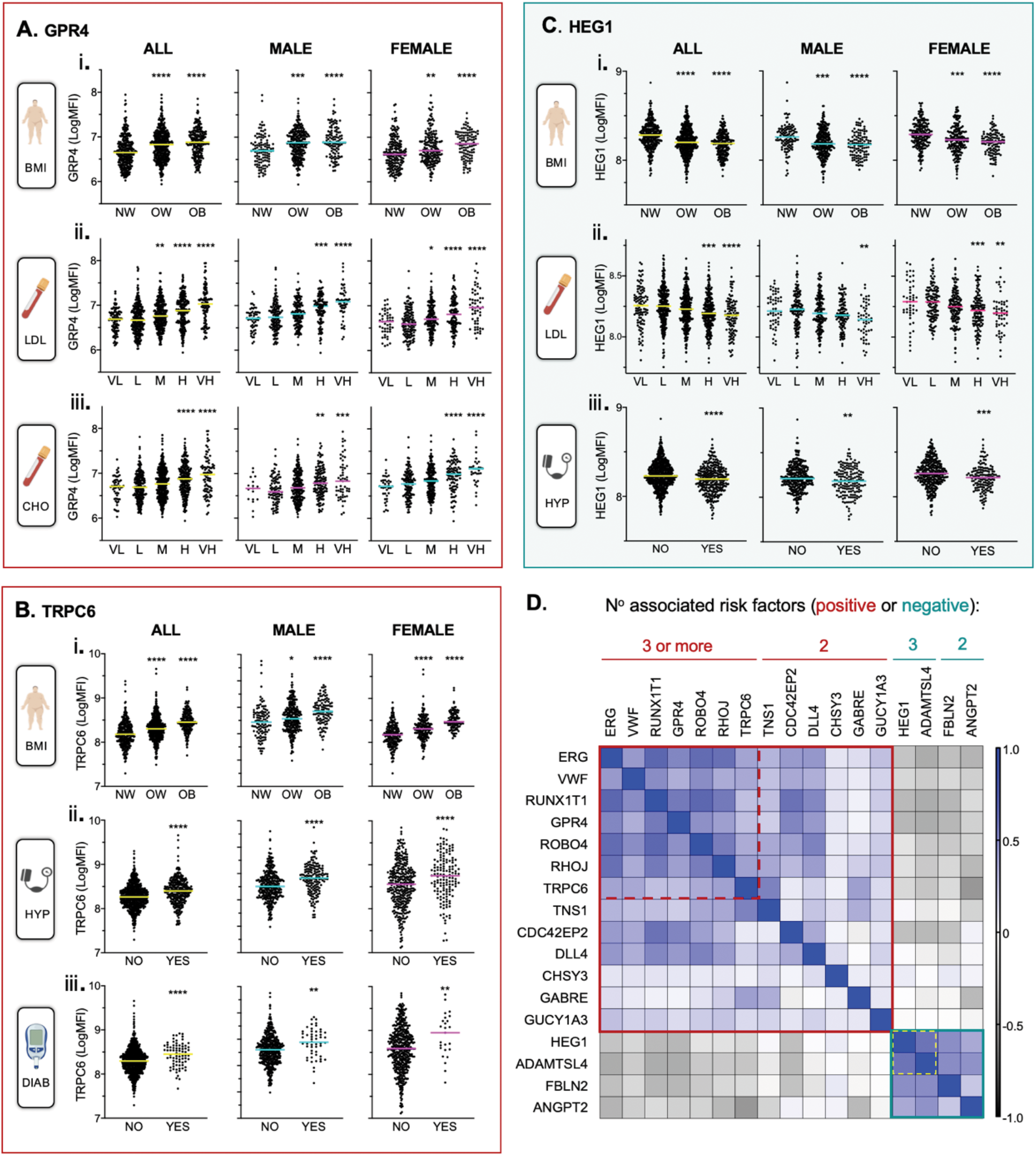
Endothelial-derived plasma protein levels can be associated with multiple cardiovascular disease (CVD) risk factors. 216 endothelial (EC)-enriched proteins were measured in plasma samples from male (n=498) and female (n=507) participants in the *Swedish CArdioPulmonary bioImage Study* (SCAPIS) pilot (n=1005). Candidate proteins associated with CVD risk factors: body mass index (BMI), total cholesterol (CHO), low density lipoprotein (LDL), hypertension (HYP), smoking (SMK) or diabetes (DIAB) were identified (age and sex adjusted linear model, [Bonferroni corrected p-value]). Illustrative plots of relative plasma levels of proteins positively: (**A**) GPR4 (**B**) TRPC6, or negatively: (**C**) HEG1, associated with CVD risk factors, in (**i**) all, (**ii**) male or (**iii**) female only samples. (**D**) Heatmap matrix showing Spearman correlation coefficients between relative plasma levels of proteins associated with 2 or more risk factors across samples. Scale on right side of heatmap. P-value *<0.05**<0.01***<0.001****<0.0001 vs. normal for BMI, and vs. low for CHO and LDL (ANOVA followed by Turkey’s multiple comparisons, or unpaired t-test [HYP, DIAB]). **BMI:** normal weight [NW: 18.5-24.9], over weight [OW: 25.0-29.9], obese [OB: >30]. **CHO:** (mmol/L): very low [VL; <4.1], low [L: 4.1-5.1], moderate [M: 5.2-6.2], high [H: 6.3-7.2], very high [VH: ≥7.3]. **LDL:** (mmol/L): very low [VL: <2.6], low [L: 2.6-3.4], moderate [M: 3.5-4.1], high [H: 4.2-4.9], very high [VH: ≥ 5].

In all of these 17 cases, variation in protein levels across risk categories were comparable between the full cohort, male only and female only samples. However, 10/13 (77%) of the EC-derived proteins that were positively associated with multiple risk factors were present at significantly lower levels in female samples overall, compared to male samples (Figure S2 A). Two of the four proteins that were negatively associated with risk factors were present at significantly higher levels in female samples, compared to male (Figure S2 B). No significant difference between the sexes was observed for the remaining proteins. Thus, overall females had lower levels of risk associated proteins and higher levels of those inversely correlated with elevated risk.

To determine if there was a potential relationship between risk associated proteins within individuals, we calculated correlation coefficients between protein levels across the full sample set. Proteins with the same effect direction, i.e., positive or negatively associated with risk factors, generally correlated with each other across the cohort (Figure 2D [Figure S3 A shows all values]). The mean correlation was strongest between proteins associated with three or more risk factors (corr. ± std. dev. 0.50 ± 0.12 [positive association], 0.62 [negative association] all p<0.00001) (Figure 2D, indicated by red and yellow dashed lines, respectively). Positively associated proteins were generally modestly inversely correlated with negatively associated ones, the strongest inverse relationship observed between those associated with 3+ risk factors (mean corr. ± std. dev. −0.27 ± 0.09 p<0.00001). Equivalent analysis in male only, or female only, sample sets generated comparable results (Figure S3 B and C). Thus, individuals with high plasma levels of any one of the multiple risk-associated EC-derived proteins tended to have high levels of other such proteins, together with lower levels of (potentially) ‘protective’ EC-derived proteins. This is consistent with the concept that a plasma biomarker panel combination could indicate degree of EC dysfunction on an individual level.

### EC-derived plasma proteins sex specifically associated with CVD risk factors

There is relatively little known about sex-specific plasma protein profiles associated with CVD risk factors. We performed a sex specific subgroup analysis (female n=507, male n=498), which identified 17 EC-derived plasma proteins associated with CVD risk factor(s) *only* in females *or* males (Figure 3) (Table S1, Tab_3) (for details on classification criteria, see methods).

**Figure 3.**
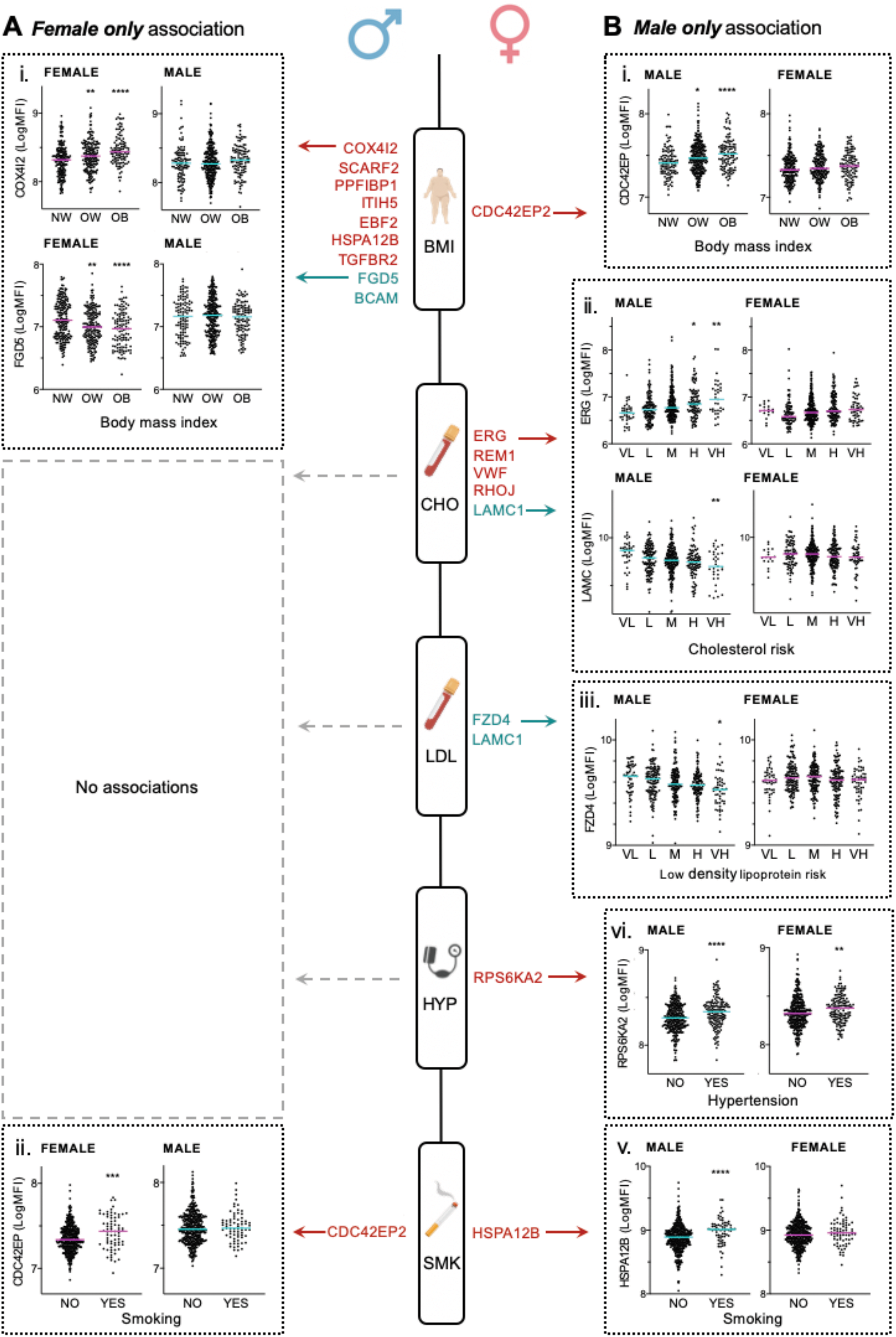
Endothelial-derived plasma protein levels can be sex-specifically associated with cardiovascular disease (CVD) risk factors. 216 endothelial (EC)-enriched proteins were measured in plasma samples from male (n=498) or female (n=507) participants in the *Swedish CArdioPulmonary bioImage Study* (SCAPIS) pilot. Candidates associated with body mass index (BMI), total cholesterol (CHO), low density lipoprotein (LDL), hypertension (HYP) or smoking (SMK) (age adjusted linear model [Bonferroni corrected p-value]) in (**A**) females only or (**B**) males only were identified. Red or green text indicates a positive or negative association, respectively, with the adjacent risk factor. Illustrative plots of plasma levels of proteins sex-specifically associated with (**i**) BMI, (**ii**) total CHO, (**iii**) LDL, (**iv**) hypertension or (**v**) smoking. *<0.05 **<0.01 ***<0.001****<0.0001 vs. normal for BMI, and vs. low for CHO and LDL (ANOVA followed by Turkey’s multiple comparisons, or unpaired t-test [HYP, DIAB]). **BMI:** normal weight [NW: 18.5-24.9], over weight [OW: 25.0-29.9], obese [OB: >30]. **CHO:** (mmol/L): very low [VL;<4.1], low [L: 4.1-5.1], moderate [M: 5.2-6.2], high [H: 6.3-7.2], very high [VH: ≥ 7.3]. **LDL:** (mmol/L): very low [VL: <2.6], low [L: 2.6-3.4], moderate [M: 3.5-4.1], high [H: 4.2-4.9], very high [VH: ≥ 5].

10 proteins were associated with CVD risk factors in females only (Figure 3A, red or green text indicates positive or negative association, respectively); of these, 9/10 (90%) were associated with BMI and 7 of these 9 (78%) (all except HSPA12B and TGFBR2) were not associated with any other risk factors in either sex, indicating a link between these proteins and a specific risk factor exposure (BMI), in a specific sex (female). Representive plots show relative plasma expression levels of COX4I2 and FGD5 (Figure 3A.i) in female vs. male samples. CDC42EP2 was positively associated with smoking in females only (Figure 3A.ii) but was also linked to exposure to other CVD risk factors, in both sexes (Figure 1B). To determine if there is a potential relationship between plasma levels of female-specific risk associated proteins, we calculated correlation coefficients across the female only sample set (Figure S4 A.i). Proteins with the same effect direction, i.e., positive or negatively associated with BMI (red or green bar on the heatmap), generally correlated across the cohort (mean corr. ± std. dev. positive association corr. 0.47 ± 0.19, negative corr. 0.78) (Figure S4 A.i). Equivalent analysis in the male only sample set (of female only risk-associated proteins) generated comparable results (Figure S4B.ii), indicating a consistent relationship between plasma levels of these proteins in both sexes, independent of changes associated with risk factor exposure. There were 9 proteins linked to a CVD risk factor in males only; 7/9 (78%) of these associations were with CHO or LDL levels (Figure 3B). Illustrative plots show expression of ERG (positively associated with CHO, Figure 3B ii), LAMC1 (negatively associated with CHO, Figure 3B ii) and FZD4 (negatively associated with LDL, Figure 3B.iii) vs. female samples. RPS6KA was more strongly associated with hypertension in males, compared to females (Figure 3B vi), although a trend was apparent in both sexes (association failed to reach Bonferroni significance in females). HSPA12B was associated with smoking in males only (Figure 3B v). Of the proteins linked to a specific CVD risk factor in males only, FZD4 was the only one not associated with any other risk factor in either sex. LAMC1 and FZD4 correlated with each other in both male (Figure S4 B.ii) and female (Figure S4 B.i) samples. Thus, sex-specific plasma profiles were predominantly linked to BMI in females and CHO in males, potentially indicating differing responses of the endothelium to these risk factors in females and males.

### Endothelial-derived plasma proteins levels are associated with Framingham Risk Score

To determine if EC-derived plasma proteins have potential utility in risk stratification, we calculated the Framingham risk scores (FRS) for each individual, as previously described (45) (4 individuals were excluded as not all data was available) and measured the association with EC-derived protein plasma levels (linear model). 27/38 (71.1%) of EC-derived proteins that were associated with CVD risk factor(s) in the whole cohort analysis, were associated with FRS (Figure 4A and B, p-values annotated in bold). Of the proteins *not* associated with CVD risk factors in any analysis (i.e. whole cohort, or single sex), 157/163 (96%) were not associated with the FRS. Relative expression of ERG, VWF, RUNX1 and RHOJ (Figure 4A, red box, higher risk associated) and ADAMTSL4 and HEG1 (Figure 4B green box, lower risk associated) are shown across groups with increasing Framingham risk score. Analysis of variance (ANOVA) F scores (indicating the degree of variation between group means) are annotated on each plot and were consistent with the relative association p values. Thus, levels of EC-derived proteins in the plasma reflect CVD risk, as measured by the Framingham risk score.

**Figure 4.**
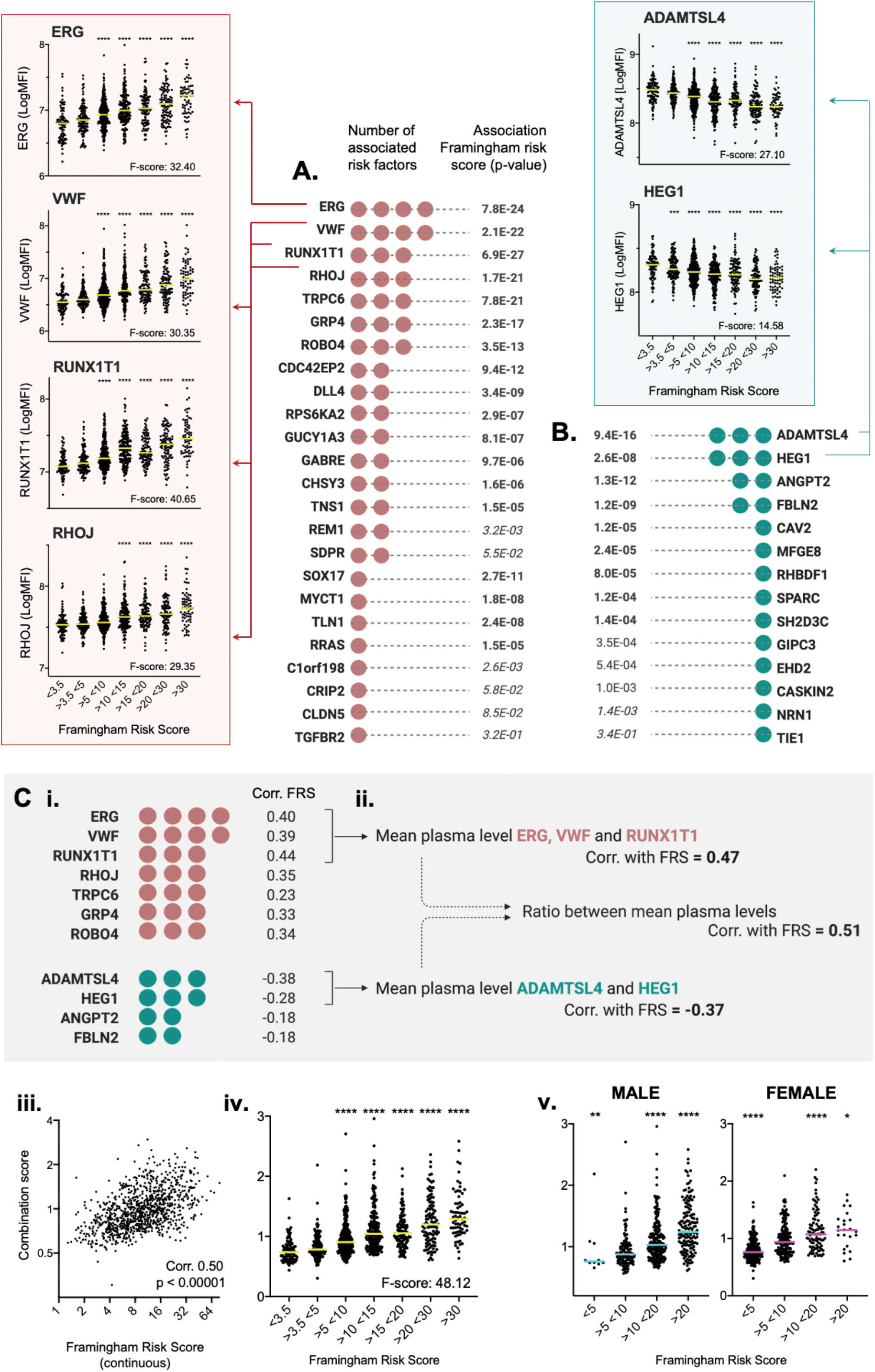
Endothelial-derived plasma protein levels are associated with Framingham risk score. 216 endothelial (EC)-enriched proteins were measured in plasma samples from male and female participants in the *Swedish CArdioPulmonary bioImage Study* (SCAPIS) pilot (n=1005). Candidates associated with body mass index (BMI), total cholesterol (CHO), low density lipoprotein (LDL), smoking, hypertension or diabetes were identified (age and sex adjusted linear model, p-value <2.0 × 10^−4^ [Bonferroni corrected]). Framingham risk scores (FRS) were calculated for each individual. Summary of EC-enriched proteins (**A**) positively or(**B**) negatively associated with one or more risk factor (adjacent red or green dots indicate number) and corresponding association between protein levels and the Framingham Risk scores across samples (linear model). Example plots in show relative protein levels in plasma across the FRS groups for proteins positively (red box) or negatively (green box) associated with FRS. (**C**) (**i**) For selected proteins, and combinations, correlation coefficients (Spearman’s) between plasma protein levels and FRS were calculated. Plots show the relationship between the EC-derived biomarker combination risk score and (**ii**) absolute FRS values, (**iii**) the FRS groups in the whole cohort and (**iv**) FRS groups in males or females only. P-value***<0.001****<0.0001 vs. group >3.5 <5 (ANOVA [F score indicates variation between group means] and Turkey’s multiple comparisons). ***Framingham risk groups***: <3.5 [n=94], >3.5 <5 [n=128], >5 <10 [n=303], >10 <15 [n=196], >15 <20 [n=107], >20 <30 [n=107], >30 [n=66].

As we observed a relationship between positively and negatively CVD risk associated EC-derived proteins (Figure 2D), we investigated the concept of using a combination of proteins for risk stratification. Spearman correlation coefficients between the FRS and proteins associated with the highest number of risk factors were calculated (Figure 4C.i) (red or green dots indicate number of positive or negative CVD risk factor associations, respectively, p-value all <1.6 × 10^−19^). Amongst those proteins positively associated with multiple CVD risk factors, ERG, VWF and RUNX1T1 showed the strongest correlation with FRS (0.40, 0.39 and 0.44, respectively). A combination value, incorporating relative levels of all three candidate proteins (mean of normalised MFI), correlated more strongly with the FRS (rho 0.47), than any individual protein (Figure 4C.ii). ADAMTSL4 and HEG1 showed the strongest FRS association amongst those proteins negatively associated with multiple CVD risk factors (−0.38 and −0.28), but an equivalent combination value (mean of normalised MFI signals) did not increase this further (corr. −0.37) (Figure 4C.ii). Finally, we calculated the ratio between these two mean values, to give a ‘combination score’; this had a higher correlation with the FRS than either combination value alone, or any individual protein (rho 0.51) (Figure 4C.ii and iii). This combination score showed a strong variation across Framingham risk score groups (F score 48.12) in the whole cohort (Figure 4C.iv), and in only male, or only female, samples (Figure 4 C.v). Thus, measurement of a panel of EC-derived proteins in the plasma more accurately reflect CVD risk, as measured by the Framingham risk score, than individual protein levels.

## DISCUSSION

Here, we identified a subset of EC-derived circulating proteins that are associated with exposure to CVD risk factors. Plasma levels of these proteins are associated with the Framingham risk score (FRS), providing proof of concept that EC-derived proteins in plasma can reflect vascular health. Accurate prediction of CVD risk is crucial when aiming for primary prevention therapies. The FRS is one of the most widely used prediction models, which is based on clinical parameters, however, it lacks individualised precision. An individual’s susceptibility to CVD based on presence of environmental CVD risk factors is determined against a background of genetic disposition, including rare variants, and history of past exposure. However, risk associated genetic variants contribute only marginally to CVD risk discrimination when incorporated into risk scores (46, 47), possibly as the contribution to life-long risk exposure does not account for the modulating effects of non-genetic risk factor exposure that varies over time. From this perspective, plasma proteomics has the advantage of integrating both genetic and environmental influences, such as life-style changes and therapeutic interventions. In particular, interrogating the proteome of one of the main players of atherosclerotic development/susceptibility, the vascular wall, holds potential to identify novel markers that can provide a direct window into the current state of pathogenesis, and also indicate new biological pathways as novel targets for therapy. Conceptually, measurement of longitudinal EC-derived proteins in plasma could provide a personalised assessment of vascular health over time, reflecting individual dynamic biological responses to dynamic changes in risk factor exposure, for example, weight loss or cessation of smoking.

One strength of our study is the use of a population-based cohort design, covering a segment of the population aged 50-64 years, where CVD risk profiles may predispose to future events, but are difficult to stratify using current methods. All participants have been evaluated and sampled in a single location, by the same personnel and procedures, thereby controlling for most of post-sampling factors that have been shown to potential bias or influence a proteomic based study (needle-to-freeze-to-analysis) (48). Another strength of our study is the targeted nature of the proteomic screening; expression specificity of a protein is a prerequisite for it to function as a useful biomarker of injury or disease of a particular tissue or cell type, e.g. plasma levels of a cardiac-specific isoform of intracellular troponin (i.e. TnT) are used to detect protein leakage from injured cardiomyocytes in myocardial infarction (49, 50) and plasma levels of prostate specific antigen can be used to screen for prostate cancer (51). In both of the aforementioned cases, which represent some of the more commonly used clinical biomarkers today, the discovery as clinical biomarker *followed* the identification of the tissue and cell specific expression patterns of the proteins. The target proteins we selected were based on our previous work identifying them as predominantly expressed in the EC compartment (37), providing highly relevant candidates to pursue as markers with potential specificity for vascular dysfunction. The Human Protein Atlas project (52) allowed us full flexibility to design such a specific screening panel that, to our knowledge, constitutes the first large EC-centric plasma analysis. Identification of such positive and negative risk associated proteins also offer starting points for investigations into function or pathways that have a potential role in the pathophysiology associated with risk factor exposure.

Other screening technologies for CVD relevant plasma proteins are available, such as commercial aptamer-based technology (www.somalogic.com) and proximity extension assays (PEA)(53). Such assays have been used to identify biomarkers associated with CVD risk factor exposure, such as blood lipids and BMI (54, 55), but these CVD screening panels are still primarily configured to detect proteins with known functions in pathophysiological processes and pathways involved in CVD (e.g. inflammation, coagulation, lipid metabolism), and thus have limited overlap with our screening panel, which has a greater focus on biomarker source, rather than previous links to CVD. Furthermore, a significant number of candidates in such pre-developed screening panels have wide tissue and/or cell type expression, which can complicate interpretation of pathophysiological relevance of identified markers. Other existing proteomics methods, such as shot-gun mass spectrometry (MS) allow for global discovery, an unbiased or agnostic interrogation of the plasma proteome that can discover completely novel targets (56). However, these techniques have a lower overall sensitivity that the affinity proteomic approach used in our study, which may bias against the detection of biologically significant, but low abundant, proteins and give less accurate quantification.

With the exception of VWF, all of the proteins we identified as positively associated with multiple CVD risk factors are normally cell-associated, rather than secreted; ERG and RUNX1T1 are transcription factors (57, 58), RHOJ and TNS1 are associated with focal adhesions (59, 60), GRP4, ROBO4, TRPC6, DLL4, GABRE are membrane proteins (61-65) and CHSY3, CDC42EP2 and GUCY1A3 are intracellular (66-68). CVD risk factor linked elevated plasma levels of these proteins could reflect EC damage, analogous to cardiac muscle troponin leakage following myocardial infarction. Conversely, proteins negatively associated with multiple CVD risk factors, HEG1, ADAMTSL4, FBLN2 and ANGPT2 are all normally secreted (69-72). Other studies have examined plasma protein association with CVD risk factor exposure and prediction using pre-designed PEA CVD panels; 102 proteins were identified as associated with baseline BMI in a study of weight loss over time, of which 88 were positively associated and 14 negatively associated (54) and 42 candidates were associated with at least one lipid fraction (triglycerides and total, LDL or HDL cholesterol) with multiple proteins overlapping groups (55). In a study of 899 participants without overt CVD in the Framingham Heart study Offspring cohort, 1129 proteins were measured, of which 156 were (positively or negatively) associated with Framingham risk score (73). However, other than VWF (74), to our knowledge there are no existing reports of the link between the proteins reported as associated in our study and CVD risk factor exposure or risk prediction, which could be explained by the limited cross over between our screening panel and those on commonly used pre-configured CVD screening panels.

There are limitations to our study. The single binder assay format used, similar to other large scale affinity proteomics assays, provides relative quantification and antibody specificity for the target protein in context of the complex matrix of plasma needs to be verified (75). As a first step in developing standardised absolute quantification assays dual binder assays, such as ELISA, or antibody-free MS-based approaches can be used for high throughput measurement of candidate proteins. Our results should then be replicated in a multicentre study. Our data could also be integerated with the genetic variation harboured by the participants to further pinpoint disease mechanism (76). The panel of proteins that we screened were originally identified as being EC-enriched under ‘normal’ conditions. Thus, it is likely that we have not measured EC-derived biomarkers that are expressed only under conditions associated with CVD risk factor exposure, e.g., inflammation. E-selectin is regulated by inflammation and highly EC-specific, and the soluble versions of this protein have been identified as a potential marker for EC-dysfunction and CVD (77). Although our knowledge of EC specific gene expression under such conditions is expanding, it is currently limited and thus challenging to perform a comprehensive screen for such candidates. In addition, we cannot rule out that the expression of the identified biomarkers is induced in cell types other than EC, as a consequence of CVD risk factor exposure. Furthermore, although we demonstrate a relationship between EC-derived plasma biomarker expression and FRS, candidate protein measurement in longitudinal samples, followed by association analysis with clinical outcomes, are needed to confirm that EC-derived plasma protein risk profile, and its variation over time, could predict development of vascular-related diseases.

In conclusion, though a targeted EC-centric analysis of plasma in relation to CVD risk factor exposure, we present the concept of using EC-derived protein profiles to infer vascular status.

## METHODS

### Samples analysed using plasma proteomics

Plasma samples were collected as part of the Swedish CArdioPulmonary BioImage Study (SCAPIS) pilot (38). Participants were sampled at the Sahlgrenska University Hospital. Whole blood was collected in EDTA anticoagulant after an overnight fast and centrifuged at 2000g for 20 minutes. Plasma aliquots were snap frozen and stored at _80°C until usage. The original SCAPIS pilot cohort contained data from 1111 individuals, however, a total of 106 of these were excluded from this study, as either (**i**) there was no blood sample available (n=43), or (ii) samples were pre-processing outliers, lacked some accompanying clinical information or were in the underweight BMI group (n=63). Thus, a total of 1005 samples were analysed. Risk factor exposure and laboratory parameters were measured as previously described (38) (also see Table S1, Tab_1).

### Antibody selection and bead-based array generation

Endothelial (EC) candidate targets proteins were selected based on our previous studies, where we identified those predominantly expressed in EC across vascular beds (37). Polyclonal antibodies, targeting 216 of these proteins, were obtained from the Human Protein Atlas project resource (www.proteinatlas.org). Plasma protein profiles were generated using affinity proteomics, as described in detail previously (78, 79). In brief, each antibody was coupled to a unique identity colour-coded magnetic beads (1.76 ug/ml, MagPlex-C, Luminex Corp.). Rabbit anti-human albumin (Dako) and donkey anti-human IgG (Jackson ImmunoResearch Laboratories) antibodies were used as controls for sample and rabbit IgG (Jackson ImmunoResearch Laboratories) and bare beads served as negative controls. Antibody-bead coupling was confirmed by R-phycoerythring-conjugated donkey anti-rabbit IgG (Jackson ImmnoResearch) prior to suspension bead array generation.

### Plasma labelling and profiling

The procedure for plasma labelling and protein profiling was performed as described previously (78, 79). Plasma samples for both cohorts were randomized according to age and sex, and distributed into microtiter plates using a liquid handling device (Freedom EVO150, Tecan). Plasma samples were diluted 1:10 in PBS and labeled with NHS-PEO4-biotin (Pierce) by liquid handling transference (CyBi-SELMA, CyBio). Labelled samples were further diluted (1:50) in assay buffer, heat-treated at 56°C for 30 minutes, cooled to RT and then combined with the suspension bead array. Unbound proteins were removed by washing and proteins captured on the beads were detected through a R-phycoerythrin-conjugated streptavidin (Invitrogen), using Flexmap 3D instruments (Luminex Corp.) Protein profiles were reported as median fluorescence intensity (MFI), corresponding to relative plasma levels of each protein candidate.

### Statistical analysis

Median fluorescence Intensity (MFI) obtained as readout on the FlexMap 3D instrument (Luminex Corp), was processed and visualized in the R statistical computing software (v 3.1.2 and 3.5.1, respectively), unless stated otherwise.

A minimum of at least 32 beads per antibody/bead region were required for inclusion in the analysis. Outlier samples were identified by robust principal component analysis and excluded from further analysis (80). MFI data was normalized by i) probabilistic quotient normalization (PQN) as accounting for any potential sample dilution effects (81) and ii) multi-dimensional MA normalization to minimize the difference amount the subgroups of the samples generated by experimental factor as multiple batches (82) Log-transformation was applied to reduce right-skewness in the proteomic data distribution. In order to identify differences in protein profiles and the association with cardiovascular disease (CVD) risk factors we applied linear regression analysis for each antibody, adjusting for age and sex, to determine association with variable of interest (e.g., body mass index, hypertension, diabetes). Analysis was performed on the whole cohort (n=1005) and on sex stratified subgroups ([female n=507, male n=498]). Protein candidates were denoted as associated to a CVD risk factor in ‘all’ when: (i) Bonferroni corrected p<0.05 in the full analysis *and* (ii) p <0.05 in *both* sex stratified subgroups. Protein candidates were denoted as associated to a CVD risk factor in ‘females only’ or ‘males only’ when: (i) Bonferroni corrected p<0.05 in one sex in the split sex analysis and (ii) p>0.05 in the other sex.

CVD risk factors were analysed as continues variables or categorized accordingly the Framingham study risk score tables. Framingham Risk Score for each subject was calculated based on the previously described formula (45), where the following information was required: age (years), sex (M/F), total cholesterol (mg/dL), high-density lipoprotein cholesterol (mg/dL), current smoking (Y/N), antihypertensive treatment (Y/N), diabetes mellitus (Y/N), and physician-acquired (clinic) SBP (mm Hg).

## Data Availability

Summary data is available in Table S1

http://dx.doi.org/10.17632/xfd584yfw5.1

## AUTHOR CONTRIBUTIONS

Conceptualisation: LMB, JO, Methodology: LMB, JO, MJI, JMS, Analysis: MJI, LK, LSR, LE, MGH, Investigation: MJI, LMB, JO, Resources: MU, JMS, GB, TR, JO, LMB, Writing – Original Draft LMB, JO; Writing – Review & Editing, All; Visualisation LMB.; Supervision LMB, JO, PD, MJI; Funding Acquisition LMB, JO.

## ACKNOWLEDGEMENTS

This work was supported by funding granted to LMB from Hjärt Lungfonden (20170759, 20170537) and the Swedish Research Council (2019-01493) and to JO from Stockholm County Council (SLL/HMT, 2017-0842/0587) and Familjen Erling Perssons Foundation. We would like to thank the team from the Translational Plasma Profiling Facility at SciLifeLab, Stockholm, for support and assistance in the generation of data for this project.

---

**Figure S1.**
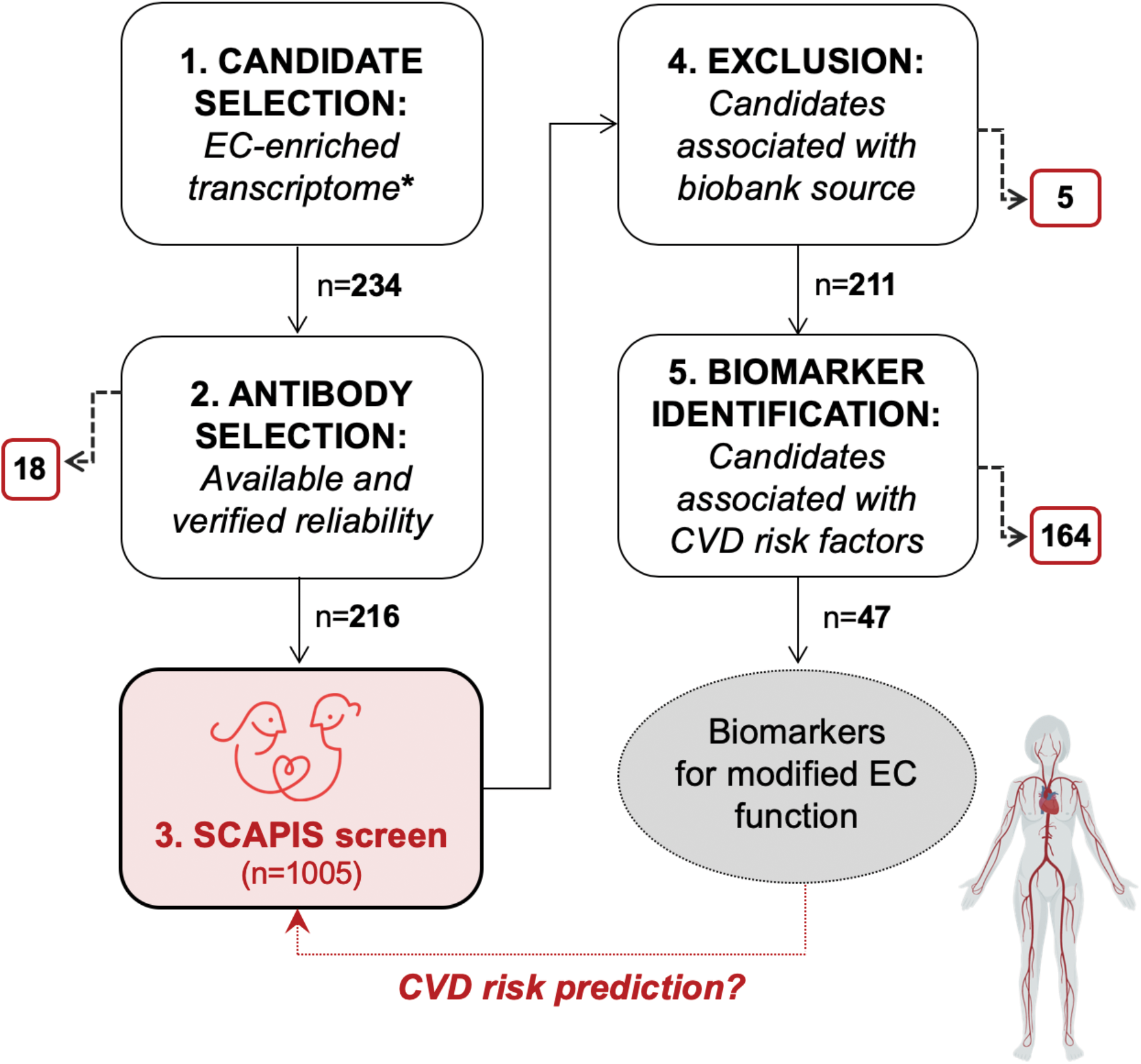
The affinity proteomic analysis workflow. (**1**) Candidate proteins for measurement in plasma were selected based on prior identification as core endothelial (EC)-enriched genes across human tissue beds. (**2**) Antibodies targeting these candidate proteins were approved for screening (n=216), or not (n=18), based on availability and reliability (see methods). (**3**) Candidate proteins were measured in plasma samples (n=1005) collected as part of the *Swedish CArdioPulmonary bioImage Study* (SCAPIS) pilot. (**4**) If the protein profile was associated with biobank source, the data was excluded (n=5). (**5**) Protein profiles that were associated (age and sex adjusted linear model, p-value <2.0 × 10^−4^ [Bonferroni corrected]) with body mass index, smoking, diagnosis of hypertension, blood low density lipoprotein, blood cholesterol or diabetes were identified (n=47) and tested for association with the Framingham risk score.

**Figure S2.**
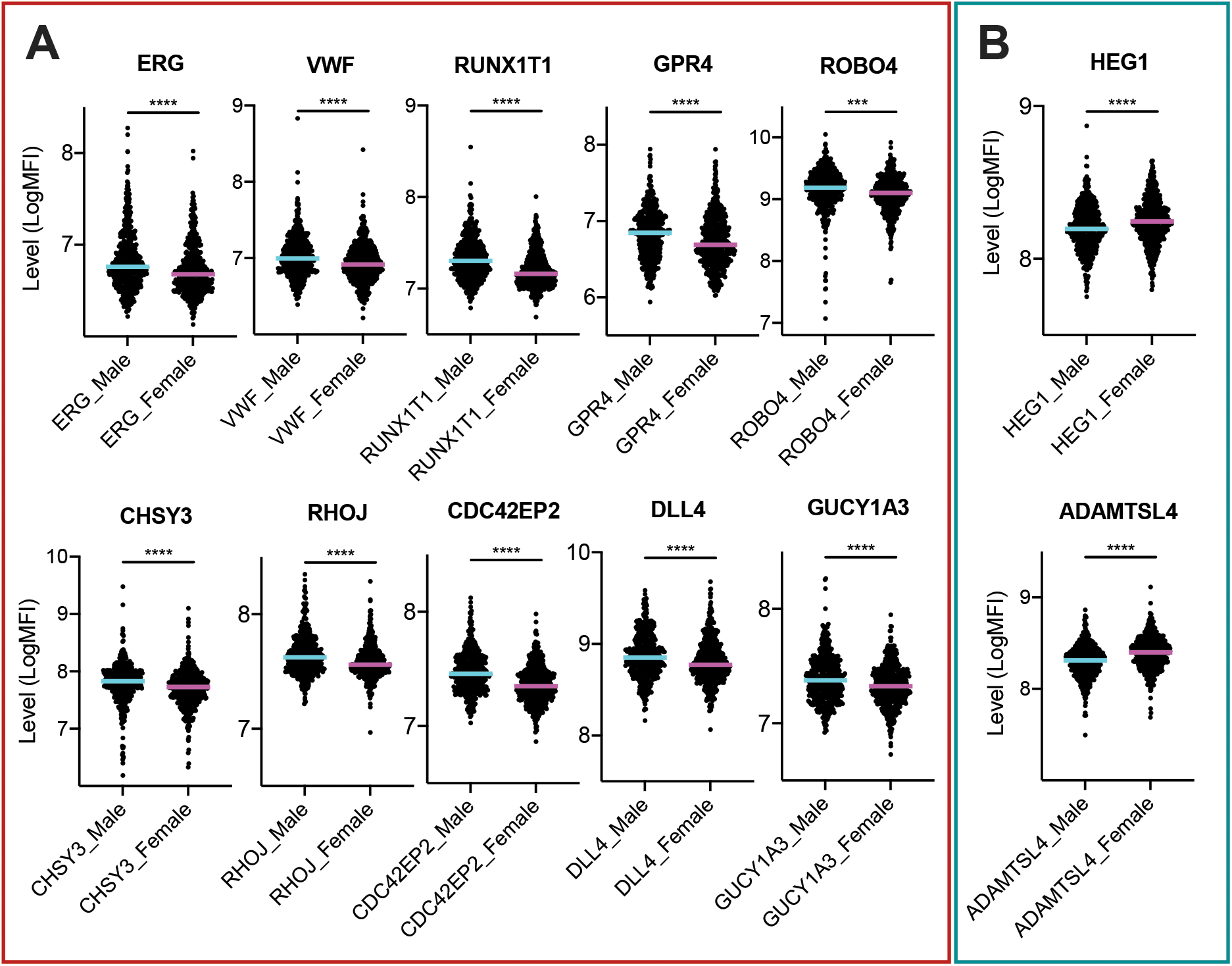
Plasma levels of endothelial-derived protein associated with multiple CVD risk factors, in male vs. female samples. Related to Figure 2. 216 endothelial (EC)-enriched proteins were measured in plasma samples from male (n=498) or female (n=507) participants in the *Swedish CArdioPulmonary bioImage Study* (SCAPIS) pilot. Relative plasma levels of proteins that were (**A**) positively (red box) or (**B**) negatively (green box) associated with multiple CVD risk factors (see Figure 2) in male and female samples. p-value *<0.05 **<0.01 ***<0.001 ****<0.0001 unpaired t-test.

**Figure S3.**
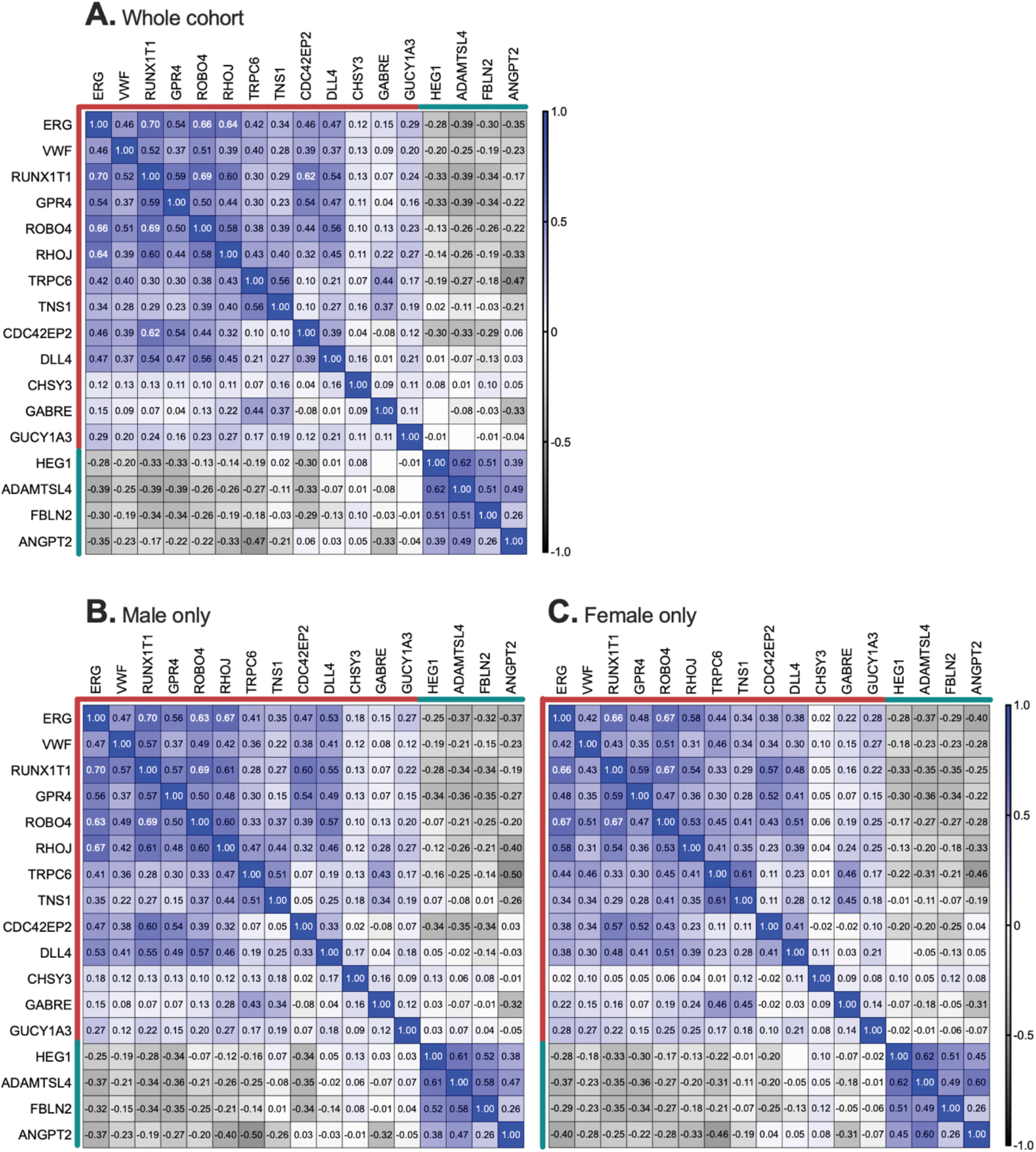
Correlation coefficients between plasma levels of endothelial-derived proteins associated with multiple CVD risk factors. Related to Figure 2. 216 endothelial (EC)-enriched proteins were measured in plasma samples from male (n=498) and female (n=507) participants in the *Swedish CArdioPulmonary bioImage Study* (SCAPIS) pilot. Heatmaps show Spearman correlation coefficients between relative plasma levels of proteins positively or negatively (indicated by red or green line, respectively) associated with multiple CVD risk factors in: (**A**) the whole cohort, (**B**) male only or (**C**) female only samples.

**Figure S4.**
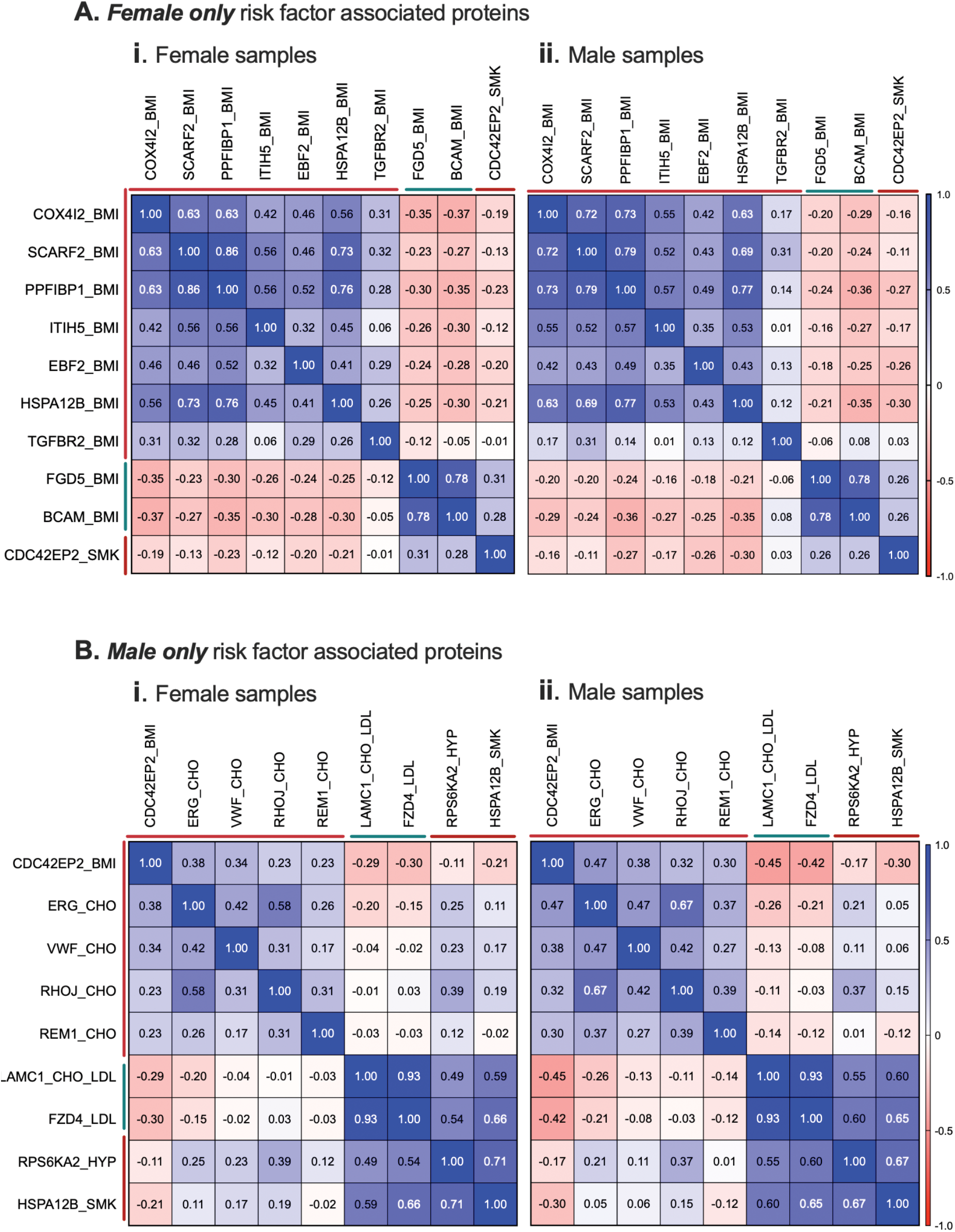
Correlation coefficients between plasma levels of endothelial-derived proteins associated with CVD risk factors, in only male or female samples. Related to Figure 4. 216 endothelial (EC)-enriched proteins were measured in plasma samples from male (n=498) or female (n=507) participants in the *Swedish CArdioPulmonary bioImage Study* (SCAPIS) pilot. Heatmaps show Spearman correlation coefficients between relative plasma levels of proteins identified as sex-specifically positively or negatively associated (indicated by red or green line, respectively) with body mass index (BMI), total cholesterol (CHO), low density lipoprotein (LDL), hypertension (HYP) or smoking (SMK).

## REFERENCES

1. J. A. Vita, Endothelial function. Circulation 124, e906–912 (2011).

2. J. S. Pober, W. C. Sessa, Evolving functions of endothelial cells in inflammation. Nat Rev Immunol 7, 803–815 (2007).

3. S. Verma, M. R. Buchanan, T. J. Anderson, Endothelial function testing as a biomarker of vascular disease. Circulation 108, 2054–2059 (2003).

4. D. S. Celermajer et al., Non-invasive detection of endothelial dysfunction in children and adults at risk of atherosclerosis. Lancet 340, 1111–1115 (1992).

5. A. Engin, Endothelial Dysfunction in Obesity. Adv Exp Med Biol 960, 345–379 (2017).

6. R. P. Brandes, Endothelial dysfunction and hypertension. Hypertension 64, 924–928 (2014).

7. S. Golbidi, L. Edvinsson, I. Laher, Smoking and Endothelial Dysfunction. Curr Vasc Pharmacol 10.2174/1573403X14666180913120015 (2018).

8. I. J. Goldberg, K. E. Bornfeldt, Lipids and the endothelium: bidirectional interactions. Curr Atheroscler Rep 15, 365 (2013).

9. M. A. Gimbrone, G. Garcia-Cardena, Endothelial Cell Dysfunction and the Pathobiology of Atherosclerosis. Circ Res 118, 620–636 (2016).

10. I. Tabas, G. Garcia-Cardena, G. K. Owens, Recent insights into the cellular biology of atherosclerosis. J Cell Biol 209, 13–22 (2015).

11. J. W. Yau, H. Teoh, S. Verma, Endothelial cell control of thrombosis. Bmc Cardiovasc Disor 15 (2015).

12. N. Mackman, New insights into the mechanisms of venous thrombosis. J Clin Invest 122, 2331–2336 (2012).

13. A. Daiber et al., Targeting vascular (endothelial) dysfunction. Br J Pharmacol 174, 1591–1619 (2017).

14. A. B. Gevaert, K. Lemmens, C. J. Vrints, E. M. Van Craenenbroeck, Targeting Endothelial Function to Treat Heart Failure with Preserved Ejection Fraction: The Promise of Exercise Training. Oxid Med Cell Longev 2017, 4865756 (2017).

15. D. Versari, E. Daghini, A. Virdis, L. Ghiadoni, S. Taddei, Endothelial Dysfunction as a Target for Prevention of Cardiovascular Disease. Diabetes Care 32, S314–S321 (2009).

16. G. M. Rubanyi, The role of endothelium in cardiovascular homeostasis and diseases. J Cardiovasc Pharmacol 22 Suppl 4, S1–14 (1993).

17. S. Verma et al., A self-fulfilling prophecy: C-reactive protein attenuates nitric oxide production and inhibits angiogenesis. Circulation 106, 913–919 (2002).

18. S. K. Venugopal, S. Devaraj, I. Yuhanna, P. Shaul, I. Jialal, Demonstration that C-reactive protein decreases eNOS expression and bioactivity in human aortic endothelial cells. Circulation 106, 1439–1441 (2002).

19. B. R. Clapp et al., Inflammation and endothelial function - Direct vascular effects of human C-reactive protein on nitric oxide bioavailability. Circulation 111, 1530–1536 (2005).

20. L. Sibal, S. C. Agarwal, P. D. Home, R. H. Boger, The Role of Asymmetric Dimethylarginine (ADMA) in Endothelial Dysfunction and Cardiovascular Disease. Curr Cardiol Rev 6, 82–90 (2010).

21. H. M. Eid, H. Arnesen, E. M. Hjerkinn, T. Lyberg, I. Seljeflot, Relationship between obesity, smoking, and the endogenous nitric oxide synthase inhibitor, asymmetric dimethylarginine. Metabolism 53, 1574–1579 (2004).

22. A. Chafai, M. F. Fromm, J. Konig, R. Maas, The prognostic biomarker L-homoarginine is a substrate of the cationic amino acid transporters CAT1, CAT2A and CAT2B. Sci Rep-Uk7 (2017).

23. J. Lew et al., Sex-Based Differences in Cardiometabolic Biomarkers. Circulation 135, 544–555 (2017).

24. J. Chen et al., Interrelationship of Multiple Endothelial Dysfunction Biomarkers with Chronic Kidney Disease. PLoS One 10, e0132047 (2015).

25. M. Budzyn et al., Plasma concentration of selected biochemical markers of endothelial dysfunction in women with various severity of chronic venous insufficiency (CVI)-A pilot study. PLoS One 13, e0191902 (2018).

26. L. F. Bittar et al., Increased Levels of Endothelial Dysfunction Markers and Matrix Metalloproteinases in Patients with Severe Post-Thrombotic Syndrome. Blood 128 (2016).

27. D. A. Scott, R. M. Palmer, The influence of tobacco smoking on adhesion molecule profiles. Tob Induc Dis 1, 7–25 (2002).

28. S. R. Patel, S. Bellary, S. Karimzad, D. Gherghel, Overweight status is associated with extensive signs of microvascular dysfunction and cardiovascular risk. Sci Rep 6, 32282 (2016).

29. C. Ferri et al., Early upregulation of endothelial adhesion molecules in obese hypertensive men. Hypertension 34, 568–573 (1999).

30. A. de la Sierra, M. Larrousse, Endothelial dysfunction is associated with increased levels of biomarkers in essential hypertension. J Hum Hypertens 24, 373–379 (2010).

31. A. D. Blann, S. K. Nadar, G. Y. Lip, The adhesion molecule P-selectin and cardiovascular disease. Eur Heart J 24, 2166–2179 (2003).

32. A. P. Arnold, L. A. Cassis, M. Eghbali, K. Reue, K. Sandberg, Sex Hormones and Sex Chromosomes Cause Sex Differences in the Development of Cardiovascular Diseases. Arterioscler Thromb Vasc Biol 37, 746–756 (2017).

33. V. Regitz-Zagrosek, G. Kararigas, Mechanistic Pathways of Sex Differences in Cardiovascular Disease. Physiol Rev 97, 1–37 (2017).

34. K. R. Kawamoto, M. B. Davis, C. S. Duvernoy, Acute Coronary Syndromes: Differences in Men and Women. Curr Atheroscler Rep 18, 73 (2016).

35. N. J. Pagidipati, E. D. Peterson, Acute coronary syndromes in women and men. Nat Rev Cardiol 13, 471–480 (2016).

36. A. J. Flammer et al., The assessment of endothelial function: from research into clinical practice. Circulation 126, 753–767 (2012).

37. L. M. Butler et al., Analysis of Body-wide Unfractionated Tissue Data to Identify a Core Human Endothelial Transcriptome. Cell Syst 3, 287–301 e283 (2016).

38. G. Bergstrom et al., The Swedish CArdioPulmonary BioImage Study: objectives and design. J Intern Med 278, 645–659 (2015).

39. S. S. Khan et al., Association of Body Mass Index With Lifetime Risk of Cardiovascular Disease and Compression of Morbidity. JAMA Cardiol 3, 280–287 (2018).

40. J. Klovaite, M. Benn, B. G. Nordestgaard, Obesity as a causal risk factor for deep venous thrombosis: a Mendelian randomization study. J Intern Med 277, 573–584 (2015).

41. P. W. Wilson et al., Prediction of first events of coronary heart disease and stroke with consideration of adiposity. Circulation 118, 124–130 (2008).

42. L. Shamai et al., Association of body mass index and lipid profiles: evaluation of a broad spectrum of body mass index patients including the morbidly obese. Obes Surg 21, 42–47 (2011).

43. H. M. Shihab et al., Body mass index and risk of incident hypertension over the life course: the Johns Hopkins Precursors Study. Circulation 126, 2983–2989 (2012).

44. V. Kotsis, S. Stabouli, S. Papakatsika, Z. Rizos, G. Parati, Mechanisms of obesity-induced hypertension. Hypertens Res 33, 386–393 (2010).

45. R. B. D’Agostino, Sr. et al., General cardiovascular risk profile for use in primary care: the Framingham Heart Study. Circulation 117, 743–753 (2008).

46. H. Holm, G. Thorleifsson, K. Stefansson, Genetic risk score and cardiovascular events in women. JAMA 303, 2032; author reply 2032-2033 (2010).

47. J. Elliott et al., Predictive Accuracy of a Polygenic Risk Score-Enhanced Prediction Model vs a Clinical Risk Score for Coronary Artery Disease. JAMA 323, 636–645 (2020).

48. U. Qundos et al., Profiling post-centrifugation delay of serum and plasma with antibody bead arrays. J Proteomics 95, 46–54 (2013).

49. K. C. Park, D. C. Gaze, P. O. Collinson, M. S. Marber, Cardiac troponins: from myocardial infarction to chronic disease. Cardiovasc Res 113, 1708–1718 (2017).

50. H. A. Katus et al., Enzyme linked immuno assay of cardiac troponin T for the detection of acute myocardial infarction in patients. J Mol Cell Cardiol 21, 1349–1353 (1989).

51. D. V. Makarov, H. B. Carter, The discovery of prostate specific antigen as a biomarker for the early detection of adenocarcinoma of the prostate. J Urol 176, 2383–2385 (2006).

52. M. Uhlen et al., Proteomics. Tissue-based map of the human proteome. Science 347, 1260419 (2015).

53. J. G. Smith, R. E. Gerszten, Emerging Affinity-Based Proteomic Technologies for Large-Scale Plasma Profiling in Cardiovascular Disease. Circulation 135, 1651–1664 (2017).

54. S. M. Figarska et al., Proteomic profiles before and during weight loss: Results from randomized trial of dietary intervention. Sci Rep 10, 7913 (2020).

55. S. M. Figarska et al., Associations of Circulating Protein Levels With Lipid Fractions in the General Population. Arterioscler Thromb Vasc Biol 38, 2505–2518 (2018).

56. X. Yin et al., Protein biomarkers of new-onset cardiovascular disease: prospective study from the systems approach to biomarker research in cardiovascular disease initiative. Arterioscler Thromb Vasc Biol 34, 939–945 (2014).

57. A. V. Shah, G. M. Birdsey, A. M. Randi, Regulation of endothelial homeostasis, vascular development and angiogenesis by the transcription factor ERG. Vascul Pharmacol 86, 3–13 (2016).

58. K. H. Liao et al., Endothelial angiogenesis is directed by RUNX1T1-regulated VEGFA, BMP4 and TGF-beta2 expression. PLoS One 12, e0179758 (2017).

59. Y. P. Shih, P. Sun, A. Wang, S. H. Lo, Tensin1 positively regulates RhoA activity through its interaction with DLC1. Biochim Biophys Acta 1853, 3258–3265 (2015).

60. L. Yuan et al., RhoJ is an endothelial cell-restricted Rho GTPase that mediates vascular morphogenesis and is regulated by the transcription factor ERG. Blood 118, 1145–1153 (2011).

61. C. A. Jones et al., Robo4 stabilizes the vascular network by inhibiting pathologic angiogenesis and endothelial hyperpermeability. Nat Med 14, 448–453 (2008).

62. E. W. Weber et al., TRPC6 is the endothelial calcium channel that regulates leukocyte transendothelial migration during the inflammatory response. J Exp Med 212, 1883–1899 (2015).

63. J. P. Liu et al., Each one of certain histidine residues in G-protein-coupled receptor GPR4 is critical for extracellular proton-induced stimulation of multiple G-protein-signaling pathways. Pharmacol Res 61, 499–505 (2010).

64. I. B. Lobov et al., Delta-like ligand 4 (Dll4) is induced by VEGF as a negative regulator of angiogenic sprouting. Proc Natl Acad Sci U S A 104, 3219–3224 (2007).

65. M. Garret et al., An mRNA encoding a putative GABA-gated chloride channel is expressed in the human cardiac conduction system. J Neurochem 68, 1382–1389 (1997).

66. A. J. Farrugia, F. Calvo, The Borg family of Cdc42 effector proteins Cdc42EP1-5. Biochem Soc Trans 44, 1709–1716 (2016).

67. T. Yada et al., Chondroitin sulfate synthase-3. Molecular cloning and characterization. J Biol Chem 278, 39711–39725 (2003).

68. E. R. Derbyshire, M. A. Marletta, Structure and regulation of soluble guanylate cyclase. Annu Rev Biochem 81, 533–559 (2012).

69. J. D. Mably, M. A. Mohideen, C. G. Burns, J. N. Chen, M. C. Fishman, heart of glass regulates the concentric growth of the heart in zebrafish. Curr Biol 13, 2138–2147 (2003).

70. L. A. Gabriel et al., ADAMTSL4, a secreted glycoprotein widely distributed in the eye, binds fibrillin-1 microfibrils and accelerates microfibril biogenesis. Invest Ophthalmol Vis Sci 53, 461–469 (2012).

71. S. L. Chapman et al., Fibulin-2 and fibulin-5 cooperatively function to form the internal elastic lamina and protect from vascular injury. Arterioscler Thromb Vasc Biol 30, 68–74 (2010).

72. R. G. Akwii, M. S. Sajib, F. T. Zahra, C. M. Mikelis, Role of Angiopoietin-2 in Vascular Physiology and Pathophysiology. Cells 8 (2019).

73. D. Ngo et al., Aptamer-Based Proteomic Profiling Reveals Novel Candidate Biomarkers and Pathways in Cardiovascular Disease. Circulation 134, 270–285 (2016).

74. B. Horvath et al., Measurement of von Willebrand factor as the marker of endothelial dysfunction in vascular diseases. Exp Clin Cardiol 9, 31–34 (2004).

75. C. Fredolini et al., Systematic assessment of antibody selectivity in plasma based on a resource of enrichment profiles. Sci Rep 9, 8324 (2019).

76. K. Suhre, M. I. McCarthy, J. M. Schwenk, Genetics meets proteomics: perspectives for large population-based studies. Nat Rev Genet 22, 19–37 (2021).

77. V. Roldan, F. Marin, G. Y. Lip, A. D. Blann, Soluble E-selectin in cardiovascular disease and its risk factors. A review of the literature. Thromb Haemost 90, 1007–1020 (2003).

78. K. Drobin, P. Nilsson, J. M. Schwenk, Highly multiplexed antibody suspension bead arrays for plasma protein profiling. Methods Mol Biol 1023, 137–145 (2013).

79. S. Bystrom et al., Affinity proteomic profiling of plasma, cerebrospinal fluid, and brain tissue within multiple sclerosis. J Proteome Res 13, 4607–4619 (2014).

80. M. Hubert, Rousseeuw, P. J., Branden, K. V., ROBPCA: A New Approach to Robust Principal Component Analysis. Technometrics 47, 64–79 (2005).

81. F. Dieterle, A. Ross, G. Schlotterbeck, H. Senn, Probabilistic quotient normalization as robust method to account for dilution of complex biological mixtures. Application in 1H NMR metabonomics. Anal Chem 78, 4281–4290 (2006).

82. M. G. Hong, W. Lee, P. Nilsson, Y. Pawitan, J. M. Schwenk, Multidimensional Normalization to Minimize Plate Effects of Suspension Bead Array Data. J Proteome Res 15, 3473–3480 (2016).

